# Association of history of metformin use with delirium and mortality: A retrospective cohort study

**DOI:** 10.1101/2022.04.03.22273209

**Authors:** Takehiko Yamanashi, Zoe-Ella EM Anderson, Manisha Modukuri, Gloria Chang, Tammy Tran, Pedro S. Marra, Nadia E. Wahba, Kaitlyn J. Crutchley, Eleanor J. Sullivan, Sydney S. Jellison, Katie R. Comp, Cade C. Akers, Alissa A. Meyer, Sangil Lee, Masaaki Iwata, Hyunkeun R. Cho, Eri Shinozaki, Gen Shinozaki

## Abstract

**Objective:** To investigate the relationship between history of metformin use and delirium risk, as well as long-term mortality.

**Methods:** In this retrospective cohort study, subjects recruited between January 2016 and March 2020 were analyzed. Logistic regression analysis was performed to investigate the relationship between metformin use and delirium. Log-rank analysis and Cox proportional hazards model were used to investigate the relationship between metformin use and 3-year mortality.

**Results:** The data from 1404 subjects were analyzed. 242 subjects were categorized into a DM-without-metformin group, and 264 subjects were categorized into a DM-with-metformin group. Prevalence of delirium was 36.0% in the DM-without-metformin group, and 29.2% in the DM-with-metformin group. A history of metformin use reduced the risk of delirium in patients with DM (OR, 0.50 [95% CI, 0.32 to 0.79]) after controlling for age, sex, and dementia status, body mass index (BMI), and insulin use. The 3-year mortality in the DM-without-metformin group (survival rate, 0.595 [95% CI, 0.512 to 0.669]) was higher than in the DM-with-metformin group (survival rate, 0.695 [95% CI, 0.604 to 0.770]) (p=0.035). A history of metformin use decreased the risk of 3-year mortality after adjustment for age, sex, Charlson Comorbidity Index, BMI, history of insulin use, and delirium status (HR, 0.69 [95% CI, 0.48 to 0.98]).

**Conclusions:** It was found that metformin usage was associated with decreased delirium prevalence and lower 3-year mortality. The potential benefit of metformin on delirium risk and mortality were shown.

## INTRODUCTION

Delirium is a severe medical illness, and its prevalence is increasing as our society ages (*1-4*). Delirium is associated with poor outcomes including extended length of stay, institutionalization after discharge from the hospital, and high mortality (*1-3, 5*). The risk of delirium increases with age, and also with medical conditions including infection and after surgery (*1-4*). At present, there is no solid understanding of the pathogenesis of delirium, and thus we do not have therapeutic or preventative methods to effectively improve care of patients with delirium. It has been shown that commonly prescribed antipsychotic medications are not helpful for treatment or prevention (*6-8*), and novel viewpoints to investigate this devastating illness are warranted.

Additional major risk factor for delirium is baseline dementia (*4*). Worse, after delirium, it is known that cognitive function further declines and dementia progression accelerates (*9*). In the recent literature, there is evidence showing that type 2 diabetes mellitus (DM) may share a key process of pathophysiology with dementia. DM and high glucose levels have been tied to increased cognitive decline and risk for dementia (*10-12*). This suggests that vascular and cellular damage induced by high blood glucose may mediate common pathological processes leading to dementia onset and progression (*13*). These evidences suggest that DM also increases the risk of delirium potentially through common underline mechanisms that increase dementia risk. However, the literature is not consistent with regard to the association between DM and delirium despite the close relationship between delirium and dementia (*14, 15*).

Of interest, among various anti-diabetic medications, it has been shown that metformin may decrease the risk of various forms of dementia, including Alzheimer’s disease (*16-19*), although the results from various studies show inconsistency (*20*). Because DM is associated with increased risk of dementia and cognitive decline, the association between anti-diabetic medication use and decreased risk of dementia was thought to be due to the better control of hyperglycemic mechanisms that could be a part of the pathogenesis of cognitive decline (*21*).

However, when metformin was compared to other anti-diabetic medications such as sulfonylureas (acetohexamide, chlorpropamide, glimepiride, glipizide, glyburide, tolazamide, and tolbutamide), the benefit for decreased risk of dementia and/or mortality was superior with metformin (*18, 19, 22-24*). To date, although multiple studies have replicated data showing that metformin seems to have benefits for decreased risk of dementia and mortality, there is very limited data about the potential role of metformin and its association with delirium, mortality, and DM.

Thus, in this report we aimed to investigate the relationship between DM and delirium risk with a focus on the influence from metformin. We hypothesized that history of metformin use is associated with lower risk for delirium. We were also interested in testing if history of metformin use can alter one of the most important patient outcomes, mortality.

## METHODS

### Design

This report is based on our previous observational cohort study of delirium at the University of Iowa Hospitals and Clinics (UIHC) (*25-29*). We conducted additional review of electronic medical records (EMRs) to gather information related to DM including body mass index (BMI), insulin use history, and metformin use history. This study was approved by the UIHC Institutional Review Board. This study complies with the Strengthening the Reporting of Observational Studies in Epidemiology reporting guideline.

### Study participants

Our previously published work describes the details of study subjects and recruitment procedures (*25-34*). Briefly, we identified eligible subjects 18 years old or older from patients at UIHC either admitted as inpatients or visiting the emergency room. Patients meeting our study inclusion criteria were approached and enrolled if they or their legally authorized representative consented. Data from a total of 1404 subjects recruited for our previous study (*25-29*) at UIHC between January 2016 and March 2020 were analyzed.

### DM and history of metformin and insulin use

Detailed metformin and insulin use history was obtained through an EMR review by the study team. Search terms such as “diabetes mellitus”, “metformin”, and “insulin” were used. Type 1 Diabetes mellitus as well as gestational diabetes were excluded from the DM group. Subjects with no history of metformin use at the time of study enrollment were classified into the metformin negative group. Other subjects, i.e., those who were taking at the time of study enrollment or had a history of metformin use before the enrollment, were classified in the metformin positive group. BMI at the time of enrollment was recorded.

### Clinical assessment and case definition

The procedures related to clinical data collection as well as definition of delirium status have been detailed in our reports published previously (*25-34*). In brief, we reviewed hospital patient records and conducted patient interviews to collect medical history and demographic characteristics. Delirium scale instruments included the Confusion Assessment Method for Intensive Care Unit (CAM-ICU) (*35*), the Delirium Observation Screening Scale (DOSS) (*36*), and the Delirium Rating Scale—Revised-98 (DRS-R-98) (*37*). The CAM-ICU and DRS were scored at the time of enrollment. As a part of the patient’s care, nursing staff recorded the DOSS score in the patient record. We defined patients’ delirium status based on CAM-ICU positive, DRS-R-98 ≥19, or DOSS ≥3, or clinical description in medical record showing the evidence of confusion or mental status change consistent with delirium (*38*). When there were questionable cases with regard to delirium status, a board-certified consultation-liaison psychiatrist (G.S.) reviewed each case for final determination for classification.

### Assessment of mortality

All-cause mortality among the study participants were gathered from a review of medical records and obituary records as previously reported (*26, 27, 29, 30, 34*).

### Statistical analysis

The statistical software EZR was used for all statistical analyses reported here (*39*). To compare the prevalence of delirium among the non-DM group, DM-with-metformin group, and DM-without-metformin group, the Chi-square test was used. To further test relationship between delirium and metformin use history in the DM group, logistic regression analysis was performed adjusting for covariates including age, sex, BMI, insulin use history, and dementia status. Furthermore, additional logistic regression analyses were performed separately for the subjects with dementia and subjects without dementia. In this logistic regression analyses, age, sex, Charlson Comorbidity Index (CCI), BMI, and insulin use history were included as covariates. For mortality analysis, Kaplan-Meier survival curves were used to visualize presentation of time to death, and log-rank statistics were used to assess significance of differences in 3-year mortality. First, we divided subjects into the following three groups: 1) non-DM group, 2) DM-without-metformin use, and 3) DM-with-metformin use. We also made subgroups divided by sex, age, presence of dementia, and presence of delirium to make Kaplan-Meier survival curves. To obtain hazard ratios (HRs) of death up to 3 years from study enrollment, we also used Cox proportional hazard regression models controlling for age, sex, CCI, BMI, insulin use history, and metformin use history using only DM subjects. Furthermore, additional Cox proportional hazard regression models controlling for same covariates were performed separately for the subjects with dementia and subjects without dementia. In addition, we performed propensity analyses. We divided subjects into two groups; non-dementia group and dementia group. The propensity for metformin use was determined using multivariable logistic regression analysis including five covariates; age, sex, CCI score, BMI, and insulin use history. The propensity scores were used to match metformin users to non-metformin users. 120 DM-with-metformin subjects were matched to 120 DM-without-metformin in non-dementia group and 28 DM-with-metformin subjects were matched to 28 DM-without-metformin in dementia group (**Supplementary Figure 1**). The p-values for comparisons among three groups were corrected by the Holm method. P-values <0.05 were considered statistically significant.

## RESULTS

### Participant demographics

A total of 1404 patients were enrolled in this study. The average patient age was 68.6 years (Standard Deviation, SD = 13.6), 48.7% were female, and 95.7% were non-Hispanic white per self-report; 898 patients were DM-negative, and 506 patients were DM-positive (**Table 1**).

**Table 1:**
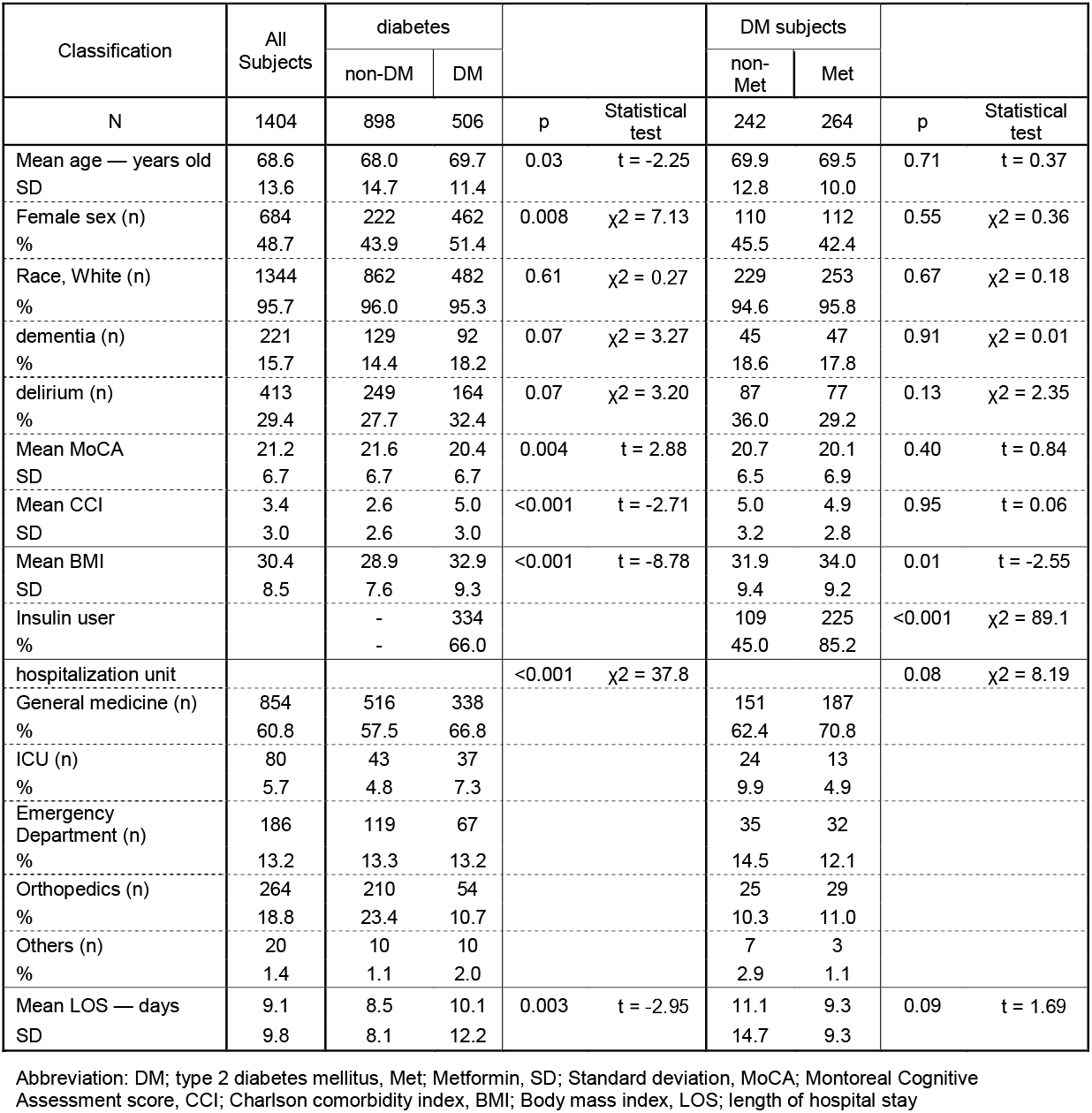
Patient Characteristics.

Among the 506 patients with DM, 264 had a history of metformin use. DM-without-metformin group had a significantly smaller BMI and less insulin use than DM-with-metformin group (mean [SD] BMI: DM-without-metformin group; 31.9 [9.4] vs DM-with-metformin group; 34.0 [9.2], t-test p = 0.01) (rate of insulin user: DM-without-metformin group; 45.0% vs DM-with-metformin group; 85.2%, chi-square test p < 0.001) (**Table 1**). Information about dementia, delirium status, Montoreal Cognitive Assessment score, CCI, hospitalization unit, and length of hospital stay are shown in Table 1.

### Delirium, DM, and history of metformin use

The prevalence of delirium in the DM-without-metformin group (36.0%) was significantly higher than it was in the non-DM group (27.7%) (p = 0.048, Chi-square test corrected by Holm method) (**Figure 1**). The prevalence of delirium in the DM-with-metformin group (29.2%) was lower than it was in the DM-without-metformin group (36.0%), but this result was not statistically significant (p = 0.25, Chi-square test corrected by Holm method) (**Figure 1**). Logistic regression analysis using only DM subjects showed that a history of metformin use reduced the risk of delirium in patients with DM (OR = 0.50, 95% CI: 0.32–0.79, p = 0.003) even after controlling for age, sex, dementia status, BMI, and history of insulin use (**Table 2**). When logistic regression analyses were performed separately for the DM subjects with dementia and DM subjects without dementia, the results showed that metformin use history reduced risk of delirium both in non-dementia group (OR: 0.54, 95%CI: 0.32–0.91, p = 0.02) (**Supplementary Table 1**) and dementia group (OR: 0.40, 95%CI: 0.13–1.21, p = 0.11) (**Supplementary Table 2**) after controlling for age, sex, CCI, BMI, and history of insulin use, although dementia group did not reach statistically significant level likely due to reduced sample size.

**Table 2:**
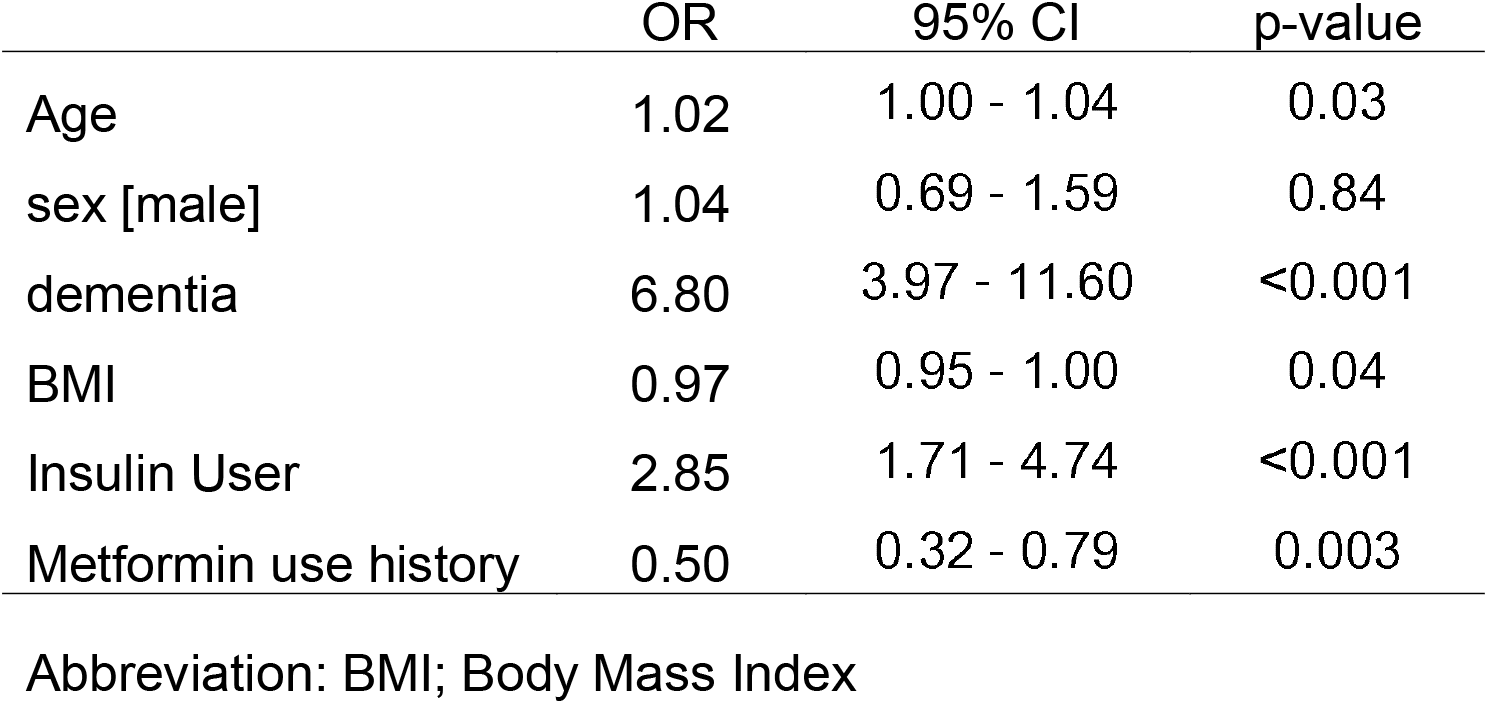
Result of the Logistic regression in subjects with diabetes (N=506)

**Figure 1.**
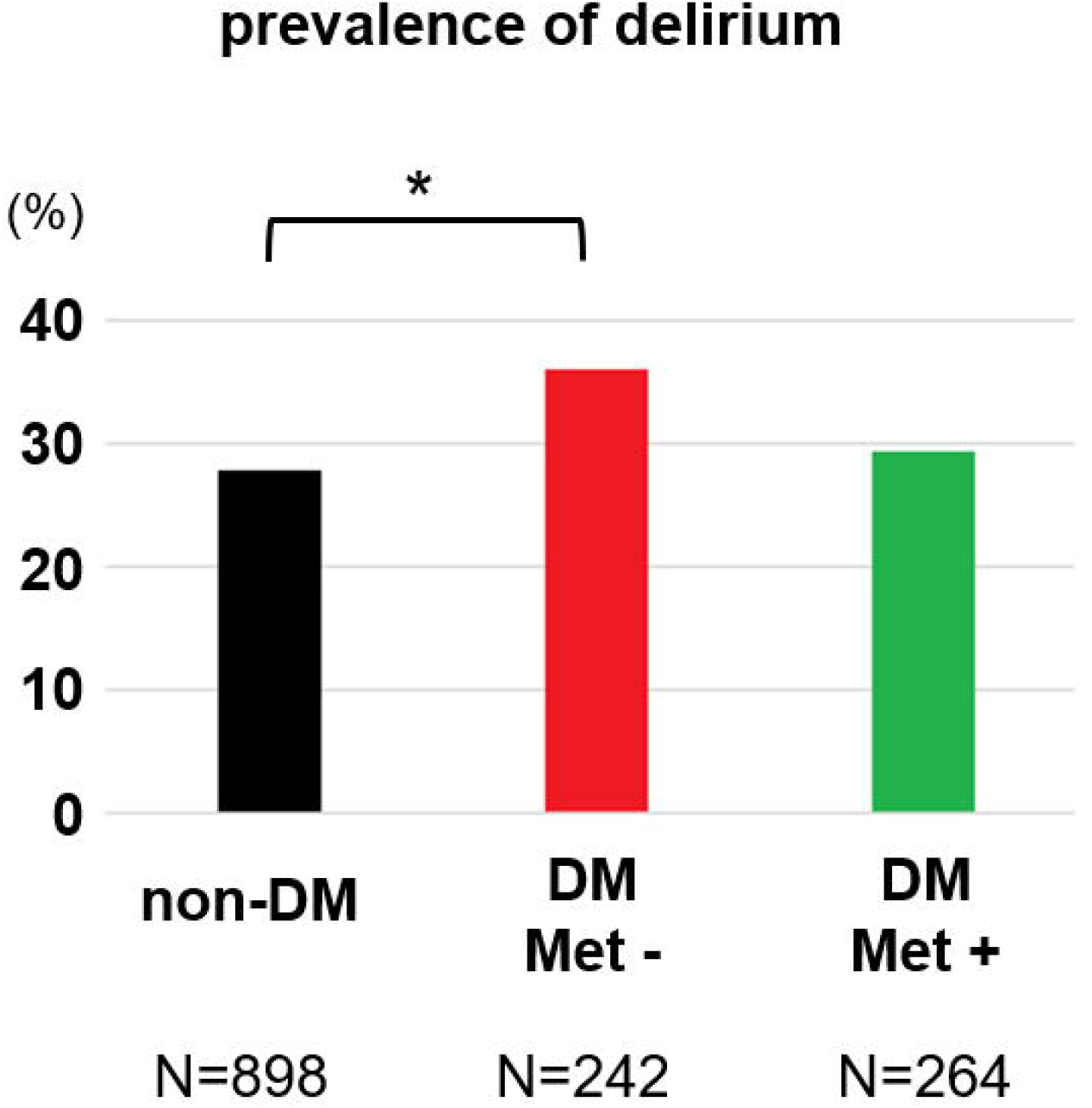
Prevalence of delirium by comparing three patient groups based on their DM status and history of metformin use. The Chi-square test corrected by Holm method showed significant difference between non-DM group and DM-without-metformin group (p=0.048).

### Mortality risk factors; benefit of history of metformin use

First, 3-year mortality was compared between the following three groups: the non-DM group, DM-without-metformin group, and DM-with-metformin group. Mortality for the DM-without-metformin group (survival rate: 0.595, 95% CI: 0.512–0.669) was significantly higher than mortality for the non-DM group (survival rate: 0.715, 95% CI: 0.672–0.753) (p = 0.0036, log-rank test corrected by Holm method). On the other hand, mortality for the DM-with-metformin group (survival rate: 0.695, 95% CI: 0.604–0.770) was significantly lower than mortality for the DM-without-metformin group (p = 0.035, log-rank test corrected by Holm method). The mortality for DM-with-metformin group is almost exactly as good as that of non-DM patients (p = 0.91, log-rank test corrected by Holm method) (**Figure 2**). The same results were replicated in younger (age < 65) subgroup, aged (age ≥ 65) subgroup, female subgroup, male subgroup, non-dementia subgroup, and non-delirium subgroup (**Supplementary Figures 2A-D, 3A, and 3C**). Intriguing differences were found in dementia subgroup and delirium subgroup. Although it did not reach statistical significance, the mortality among DM patients who have used metformin were lower even when compared to that of non-DM group (**Supplementary Figure 3B and 3D**). Cox proportional hazard model showed that history of metformin use significantly decreased risk of 3-year mortality after adjustment for age, sex, CCI, BMI, history of insulin use, and delirium status (HR = 0.69, 95%CI: 0.48–0.98, p = 0.038) (**Table 3**). When cox proportional hazard models were performed separately for the DM subjects with dementia and DM subjects without dementia, the results showed that metformin use history did not reduce risk of mortality in non-dementia group (HR: 0.89, 95%CI: 0.59–1.35, p = 0.59) (**Supplementary Table 3**), but metformin exposure reduced risk of mortality among dementia group (HR: 0.40, 95%CI: 0.18– 0.87, p = 0.02) (**Supplementary Table 4**). The GLOBAL tests for the proportional hazards assumptions were not statistically significant (**Table 3, Supplementary Table 3, and Supplementary Table 4**).

**Table 3:** Result of the Cox proportional hazard model in subjects with diabetes (N=506)

|  | HR | 95% CI | p-value |
| --- | --- | --- | --- |
| Age | 1.05 | 1.03 - 1.07 | <0.001 |
| sex male | 1.37 | 0.97 - 1.92 | 0.07 |
| CCI | 1.15 | 1.10 - 1.20 | <0.001 |
| BMI | 1.01 | 0.99 - 1.03 | 0.35 |
| Insulin User | 0.91 | 0.63 - 1.32 | 0.62 |
| delirium | 1.55 | 1.10 - 2.18 | 0.01 |
| Metformin use<br>history | 0.69 | 0.48 - 0.98 | 0.04 |

Abbreviation: CCI; Charlson Comorbidity Index, BMI; Body Mass Index
Likelihood ratio test: $p < 0.001$
Wald test: $p < 0.001$
Score (logrank) test: $p < 0.001$
GLOBAL test for the proportional hazards assumption: $p = 0.14$

**Figure 2.**
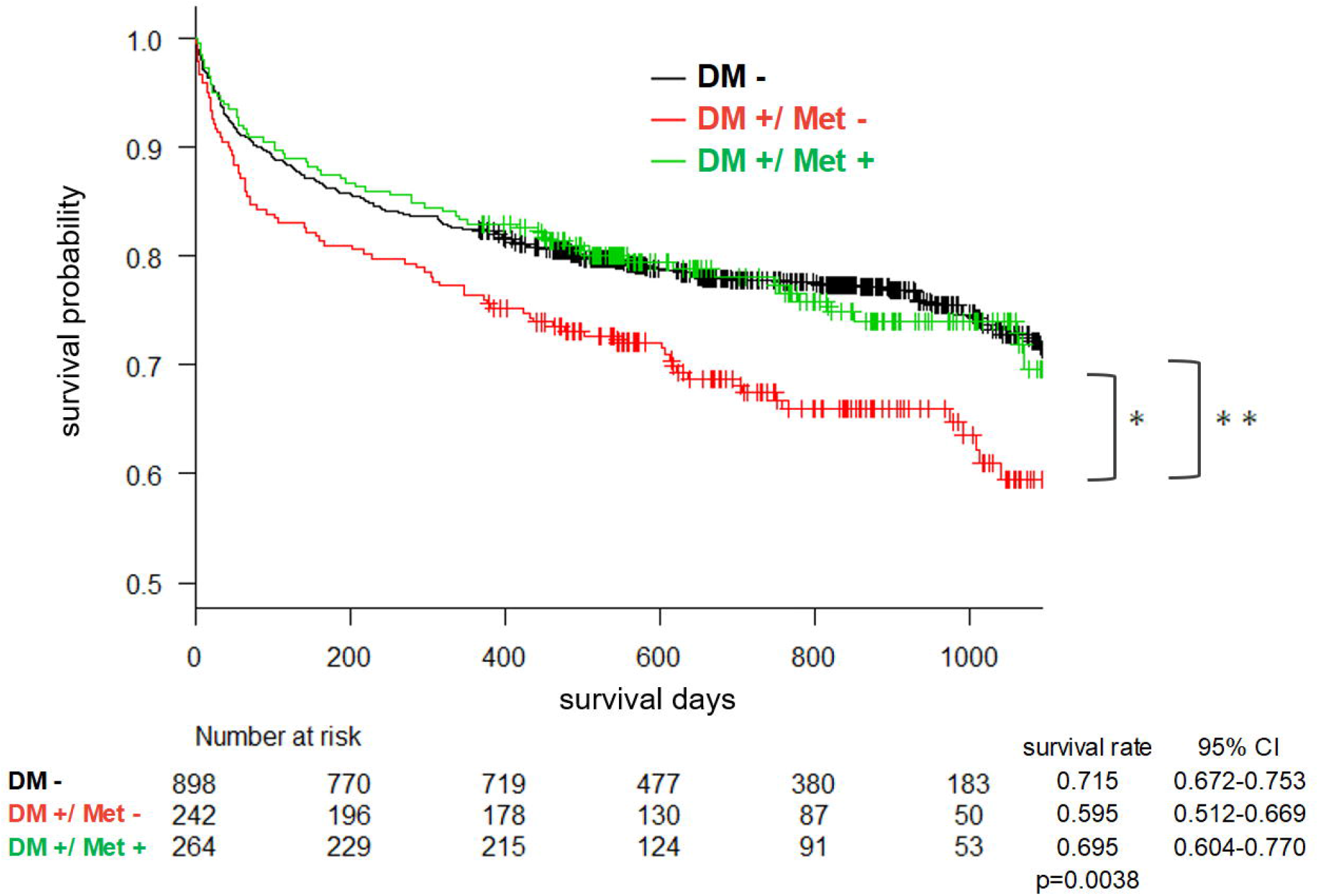
Kaplan-Meier cumulative survival curve over 3 years based on the three-group comparison.

### Results of propensity analyses

The characteristics of matched subjects are shown in **Supplementary Table 5**. The prevalence of delirium in the DM-with-metformin group was lower than one in the DM-without-metformin group both in non-dementia matched group (DM-without-metformin group: 37.5% vs DM-with-metformin group: 25.8%, Chi-square test p=0.07) and dementia matched group (DM-without-metformin group; 71.4% vs DM-with-metformin group; 60.7%, Chi-square test p=0.57), but without statistical significance (**Supplementary Table 5**). Logistic regression analysis showed a reduced risk of delirium by a history of metformin use both in non-dementia matched group (OR = 0.58, 95% CI: 0.34–1.01, p = 0.053) and dementia matched group (OR = 0.62, 95% CI: 0.20– 1.89, p = 0.40), but without statistical significance. Mortality for the DM-with-metformin group was significantly lower than mortality for the DM-without-metformin group both in non-dementia matched group (DM-without-metformin group; survival rate: 0.595, 95% CI: 0.476–0.700 vs DM-with-metformin group; survival rate: 0.658, 95% CI: 0.536–0.755, log-rank p = 0.29) and dementia matched group (DM-without-metformin group; survival rate: 0.382, 95% CI: 0.147– 0.617 vs DM-with-metformin group; survival rate: 0.709, 95% CI: 0.476–0.853, log-rank p = 0.07), but with statistical significance (**Supplementary Figure 4**). Cox proportional hazards model showed a reduced risk of mortality by a history of metformin use both in non-dementia matched group (HR = 0.78, 95% CI: 0.50–1.24, p = 0.29) and dementia matched group (OR = 0.44, 95% CI: 0.18–1.11, p = 0.08), but without statistical significance.

## DISCUSSION

This large-cohort study examined whether history of DM as well as metformin use are associated with delirium. Another investigation was if DM and history of metformin use are altering mortality risk. In the data presented here, higher prevalence of delirium and increased mortality were observed in DM patients without a history of metformin use compared to non-DM patients. On the other hand, DM patients with a history of metformin use showed lower prevalence of delirium in our data, and this could be a reason why past literature investigating relationship between DM and delirium was inconclusive, as most likely those subjects categorized with DM included those who were on metformin, and thus had less prevalence of delirium in average over DM patients with and without metformin use. Additionally, we found that a history of metformin use was associated with decreased risk of delirium and mortality even after adjusting for many potential confounding variables. Our data indicate the potential positive benefit of metformin on delirium risk and mortality, providing insights about additional pathophysiological mechanisms of delirium and potential therapeutic and preventative opportunities for this devastating illness commonly seen among the aged population. However, it still remains unclear whether diabetes increases the risk of delirium or mortality as we have not analyzed the direct effect of diabetes on the risk of delirium and mortality with prospective clinical study including various important confounding factors.

The association observed here in this report between DM and delirium itself is not new, although previous data were mixed (*14, 15*). However, the preventative role of metformin related to delirium is unique, and this is the first report showing such potential relationship. There has been little study investigating relationship between metformin and delirium. One study, which used the U.S. Food and Drug Administration adverse events reporting system (FAERS), reported metformin as a potential delirium-inducing drug, contrary to our result (*40*). The discrepancy in these results might be due to methodological difference because the data in the FAERS are not designed to specifically investigate relationship between metformin and delirium but rather screening of large variety of commonly prescribed medications (*40*). To solve this inconsistency, a future randomized clinical trial to test metformin for its effect on delirium risk would be of importance.

In addition, metformin has been reported to be beneficial for survival among various patient populations including cancer patients (*41-43*). Our data presented here showed its potential benefit for survival regardless of delirium status. It is of importance to note that DM patients without a history of metformin use showing higher mortality was not simply due to their having a severe form of DM, because subjects in this group are defined as those who have never been on metformin, and they are not the group that was diagnosed with DM, treated with metformin first, and was switched to insulin due to their poor control of DM while they were on metformin. In fact, in our data set, BMI and insulin use ratio were lower in DM-without-metformin group than ones in DM-with-metformin group, suggesting that DM-without-metformin group had potentially less severe diabetes.

Basic research studies have shown that metformin appears to target a number of aging-related mechanisms (*44, 45*). It was reported that metformin influenced pro-inflammatory cytokines such as IL-6 and TNF-alpha to suppress inflammation through modulating the NF-kB pathway (*46, 47*). Furthermore, metformin was reported to activate AMPK and inhibits mTOR (*48, 49*), which is known to influence the aging process (*45, 49, 50*). It is possible that these effects by metformin on inflammation and pathways involving AMPK and mTOR can decrease delirium risk and prolong patient’s life, although evidence for these effects in humans are limited.

Our data showed this potential benefit of metformin. The question is whether people without DM should start taking metformin. It is an important question that should be carefully explored, as metformin can have problematic side effects including vitamin B12 deficiency if taken for the long term (*51*). Then the next question is if patients at risk for delirium such as those going through major surgery (e.g., cardiac, orthopedic, or neurosurgery) should take metformin even for a short period of time preoperatively and postoperatively. This needs to be answered by future clinical trials, and we believe these are worth conducting to improve our patient care.

## LIMITATIONS

We acknowledge the following limitations in this report. First, because history of metformin use was obtained from hospital records by retrospective chart review, it is possible to include false-positive cases when patients were prescribed with metformin but did not take it because of intolerable side effects or non-adherence to it, and false-negative situations such as metformin prescribed by other providers outside of our hospital network. Second, the definition of delirium is not based on the gold standard psychiatric assessment based on DSM-5 criteria (*52*). However, our categorization methods have been effective enough to show discrete mortality risk based on our clinical classification of delirium as shown in our previous reports (*25-29*). Also, Inouye et al. reported that retrospective chart review can capture delirium with reasonably good sensitivity and specificity (*38*). Third, the definition of DM is based on chart review as well, although our classification of DM clearly differentiates mortality risk, supporting the validity of our approach. Fourth, we did not control for metformin dose or duration. Total dose information might be important because duration or dose of metformin can alter its effect, as a previous animal study showed (*53*). Fifth, other anti-diabetic medication other than insulin were not recorded and incorporated in our investigation, as it has been repeatedly reported that when compared to metformin, other anti-diabetic medications did not show benefit for mortality (*22-24*). Lastly, our data does not necessarily show causal relationship of metformin use and risk for delirium or mortality. However, metformin was used prior to the occurrence of delirium as well as death, suggesting a strong possibility of a beneficial effect from metformin in decreasing risk for delirium and increasing chance of survival. To address this question more precisely, prospective clinical trials are needed. Despite all these potential limitations, we found significant associations among metformin, DM, and delirium, as well as all-cause mortality.

## CONCLUSION

In this report, we showed the potential benefit of metformin in decreasing the risk of delirium and mortality.

## Data Availability

All data produced in the present study are available upon reasonable request to the authors

## Funding / Support

None.

## Declaration of Interest

Gen Shinozaki is co-founder of Predelix Medical LLC and has pending patents as follows: “Non-invasive device for predicting and screening delirium”, PCT application no. PCT/US2016/064937 and US provisional patent no. 62/263,325; “Prediction of patient outcomes with a novel electroencephalography device”, US provisional patent no. 62/829,411; “Epigenetic Biomarker of Delirium Risk” in the PCT Application No. PCT/US19/51276, and in U.S. Provisional Patent No. 62/731,599. All other authors have declared that no conflict of interest exists.

## Acknowledgment

The authors thank the patients who participated in this study. Also, we would like to thank Paul J. Casella (University of Iowa Carver College of Medicine) for English language editing.

## Author Contributions

T.Y. organized and analyzed data, and edited of the final form of the manuscript. Z.E.A., M.M., G.C., and T.T. collected the clinical data, organized it and analyzed for initial preparation, and wrote the initial draft of the manuscript. P.S.M., N.E.W., K.J.C., E.J.S., S.S.J., K.R.C., C.C.A., and A.A.M. collected the clinical data. S.L. and E.S. organized data collection process and critically reviewed the manuscript. M.I. critically reviewed the manuscript. H.R.C. analyzed data. G.S. conceived ideas of the study, planned its design and coordination, wrote the initial draft and edited the final form of the manuscript.

## Access to Data and Data Analysis

G.S. had full access to all the data in the study and takes responsibility for the integrity of the data and the accuracy of the data analysis

## Data Availability

The data that support the findings of this study are available from the corresponding author, G.S., upon reasonable request.

## Notes

### Funding Statement

This study did not receive any funding

### Author Declarations

IRB of the University of Iowa Hospitals and Clinics gave ethical approval for this work.

## References

1. S. K. Inouye, Delirium in older persons. N. Engl. J. Med. 354, 1157–1165 (2006).

2. T. G. Fong, S. R. Tulebaev, S. K. Inouye, Delirium in elderly adults: diagnosis, prevention and treatment. Nat. Rev. Neurol. 5, 210–220 (2009).

3. S. K. Inouye, R. G. Westendorp, J. S. Saczynski, Delirium in elderly people. Lancet 383, 911–922 (2014).

4. J. R. Maldonado, Delirium pathophysiology: An updated hypothesis of the etiology of acute brain failure. Int. J. Geriatr. Psychiatry 33, 1428–1457 (2018).

5. J. McCusker, M. Cole, M. Abrahamowicz, F. Primeau, E. Belzile, Delirium predicts 12-month mortality. Arch. Intern. Med. 162, 457–463 (2002).

6. T. D. Girard et al., Haloperidol and Ziprasidone for Treatment of Delirium in Critical Illness. N. Engl. J. Med. 379, 2506–2516 (2018).

7. E. S. Oh et al., Antipsychotics for Preventing Delirium in Hospitalized Adults: A Systematic Review. Ann Intern Med 171, 474–484 (2019).

8. R. Nikooie et al., Antipsychotics for Treating Delirium in Hospitalized Adults: A Systematic Review. Ann. Intern. Med. 171, 485–495 (2019).

9. P. P. Pandharipande et al., Long-term cognitive impairment after critical illness. N. Engl. J. Med. 369, 1306–1316 (2013).

10. G. Cheng, C. Huang, H. Deng, H. Wang, Diabetes as a risk factor for dementia and mild cognitive impairment: a meta-analysis of longitudinal studies. Intern Med J 42, 484–491 (2012).

11. P. K. Crane et al., Glucose levels and risk of dementia. N Engl J Med 369, 540–548 (2013).

12. C. Barbiellini Amidei et al., Association Between Age at Diabetes Onset and Subsequent Risk of Dementia. Jama 325, 1640–1649 (2021).

13. K. Govindpani et al., Vascular Dysfunction in Alzheimer’s Disease: A Prelude to the Pathological Process or a Consequence of It? J Clin Med 8, (2019).

14. K. van Keulen et al., Diabetes and Glucose Dysregulation and Transition to Delirium in ICU Patients. Crit Care Med 46, 1444–1449 (2018).

15. S. Grover, A. Avasthi, Clinical Practice Guidelines for Management of Delirium in Elderly. Indian J Psychiatry 60, S329–s340 (2018).

16. C. P. Wang, C. Lorenzo, S. L. Habib, B. Jo, S. E. Espinoza, Differential effects of metformin on age related comorbidities in older men with type 2 diabetes. J Diabetes Complications 31, 679–686 (2017).

17. S. A. Farr et al., Metformin Improves Learning and Memory in the SAMP8 Mouse Model of Alzheimer’s Disease. J Alzheimers Dis 68, 1699–1710 (2019).

18. A. R. Orkaby, K. Cho, J. Cormack, D. R. Gagnon, J. A. Driver, Metformin vs sulfonylurea use and risk of dementia in US veterans aged ≥65 years with diabetes. Neurology 89, 1877–1885 (2017).

19. J. F. Scherrer et al., Association Between Metformin Initiation and Incident Dementia Among African American and White Veterans Health Administration Patients. Ann Fam Med 17, 352–362 (2019).

20. F. Ping, N. Jiang, Y. Li, Association between metformin and neurodegenerative diseases of observational studies: systematic review and meta-analysis. BMJ Open Diabetes Res Care 8, (2020).

21. I. K. Wium-Andersen, M. Osler, M. B. Jørgensen, J. Rungby, M. K. Wium-Andersen, Antidiabetic medication and risk of dementia in patients with type 2 diabetes: a nested case-control study. Eur. J. Endocrinol. 181, 499–507 (2019).

22. Effect of intensive blood-glucose control with metformin on complications in overweight patients with type 2 diabetes (UKPDS 34). UK Prospective Diabetes Study (UKPDS) Group. Lancet 352, 854–865 (1998).

23. J. A. Johnson, S. R. Majumdar, S. H. Simpson, E. L. Toth, Decreased mortality associated with the use of metformin compared with sulfonylurea monotherapy in type 2 diabetes. Diabetes Care 25, 2244–2248 (2002).

24. E. H. Cho, K. Han, B. Kim, D. H. Lee, Gliclazide monotherapy increases risks of all-cause mortality and has similar risk of acute myocardial infarction and stroke with glimepiride monotherapy in Korean type 2 diabetes mellitus. Medicine (Baltimore) 99, e21236 (2020).

25. G. Shinozaki et al., Delirium detection by a novel bispectral electroencephalography device in general hospital. Psychiatry Clin. Neurosci. 72, 856–863 (2018).

26. G. Shinozaki et al., Identification of Patients With High Mortality Risk and Prediction of Outcomes in Delirium by Bispectral EEG. J. Clin. Psychiatry 80, (2019).

27. T. Yamanashi et al., New Cutoff Scores for Delirium Screening Tools to Predict Patient Mortality. J. Am. Geriatr. Soc. 69, 140–147 (2021).

28. T. Yamanashi et al., Topological data analysis (TDA) enhances bispectral EEG (BSEEG) algorithm for detection of delirium. Sci. Rep. 11, 304 (2021).

29. T. Saito et al., Mortality prediction by bispectral electroencephalography among 502 patients: its role in dementia. Brain Communications, (2021).

30. T. Yamanashi et al., Mortality among patients with sepsis associated with a bispectral electroencephalography (BSEEG) score. Sci. Rep. 11, 14211 (2021).

31. T. Saito et al., Epigenetics of neuroinflammation: Immune response, inflammatory response and cholinergic synaptic involvement evidenced by genome-wide DNA methylation analysis of delirious inpatients. J. Psychiatr. Res. 129, 61–65 (2020).

32. T. Saito et al., The relationship between DNA methylation in neurotrophic genes and age as evidenced from three independent cohorts: differences by delirium status. Neurobiol. Aging 94, 227–235 (2020).

33. T. Yamanashi et al., DNA methylation in the TNF-alpha gene decreases along with aging among delirium inpatients. Neurobiol. Aging, (2021).

34. T. Yamanashi et al., Evaluation of point-of-care thumb-size bispectral electroencephalography device to quantify delirium severity and predict mortality. The British Journal of Psychiatry, 1–8 (2021).

35. E. W. Ely et al., Delirium in mechanically ventilated patients: validity and reliability of the confusion assessment method for the intensive care unit (CAM-ICU). JAMA 286, 2703–2710 (2001).

36. M. J. Schuurmans, L. M. Shortridge-Baggett, S. A. Duursma, The Delirium Observation Screening Scale: a screening instrument for delirium. Res Theory Nurs Pract 17, 31–50 (2003).

37. P. T. Trzepacz et al., Validation of the Delirium Rating Scale-revised-98: comparison with the delirium rating scale and the cognitive test for delirium. J. Neuropsychiatry Clin. Neurosci. 13, 229–242 (2001).

38. S. K. Inouye et al., A chart-based method for identification of delirium: validation compared with interviewer ratings using the confusion assessment method. J. Am. Geriatr. Soc. 53, 312–318 (2005).

39. Y. Kanda, Investigation of the freely available easy-to-use software ‘EZR’ for medical statistics. Bone Marrow Transplant. 48, 452–458 (2013).

40. A. Wong et al., Drug-Associated Delirium Identified in The Food and Drug Administration Adverse Events Reporting System. Psychological Disorders and Research, 1–7 (2019).

41. G. W. Landman et al., Metformin associated with lower cancer mortality in type 2 diabetes: ZODIAC-16. Diabetes Care 33, 322–326 (2010).

42. S. Bo et al., Cancer mortality reduction and metformin: a retrospective cohort study in type 2 diabetic patients. Diabetes Obes. Metab. 14, 23–29 (2012).

43. C. A. Bannister et al., Can people with type 2 diabetes live longer than those without? A comparison of mortality in people initiated with metformin or sulphonylurea monotherapy and matched, non-diabetic controls. Diabetes Obes. Metab. 16, 1165–1173 (2014).

44. A. S. Kulkarni, S. Gubbi, N. Barzilai, Benefits of Metformin in Attenuating the Hallmarks of Aging. Cell Metab. 32, 15–30 (2020).

45. N. Barzilai, J. P. Crandall, S. B. Kritchevsky, M. A. Espeland, Metformin as a Tool to Target Aging. Cell Metab 23, 1060–1065 (2016).

46. Z. Zhou et al., Metformin Inhibits Advanced Glycation End Products-Induced Inflammatory Response in Murine Macrophages Partly through AMPK Activation and RAGE/NFκB Pathway Suppression. J Diabetes Res 2016, 4847812 (2016).

47. A. R. Cameron et al., Anti-Inflammatory Effects of Metformin Irrespective of Diabetes Status. Circ. Res. 119, 652–665 (2016).

48. F. Yang et al., Metformin Inhibits the NLRP3 Inflammasome via AMPK/mTOR-dependent Effects in Diabetic Cardiomyopathy. Int. J. Biol. Sci. 15, 1010–1019 (2019).

49. X. Feng et al., Metformin attenuates cartilage degeneration in an experimental osteoarthritis model by regulating AMPK/mTOR. Aging (Albany N. Y.) 12, 1087–1103 (2020).

50. H. H. Glossmann, O. M. D. Lutz, Metformin and Aging: A Review. Gerontology 65, 581–590 (2019).

51. V. R. Aroda et al., Long-term Metformin Use and Vitamin B12 Deficiency in the Diabetes Prevention Program Outcomes Study. J. Clin. Endocrinol. Metab. 101, 1754–1761 (2016).

52. Diagnostic and statistical manual of mental disorders. (American Psychiatric Association, Arlington, ed. 5th, 2013).

53. W. H. Oliveira et al., Effects of metformin on inflammation and short-term memory in streptozotocin-induced diabetic mice. Brain Res. 1644, 149–160 (2016).

