## Supplementary Figures for "Association of history of metformin use with delirium and mortality: A retrospective cohort study"

**Supplementary Figure 1**

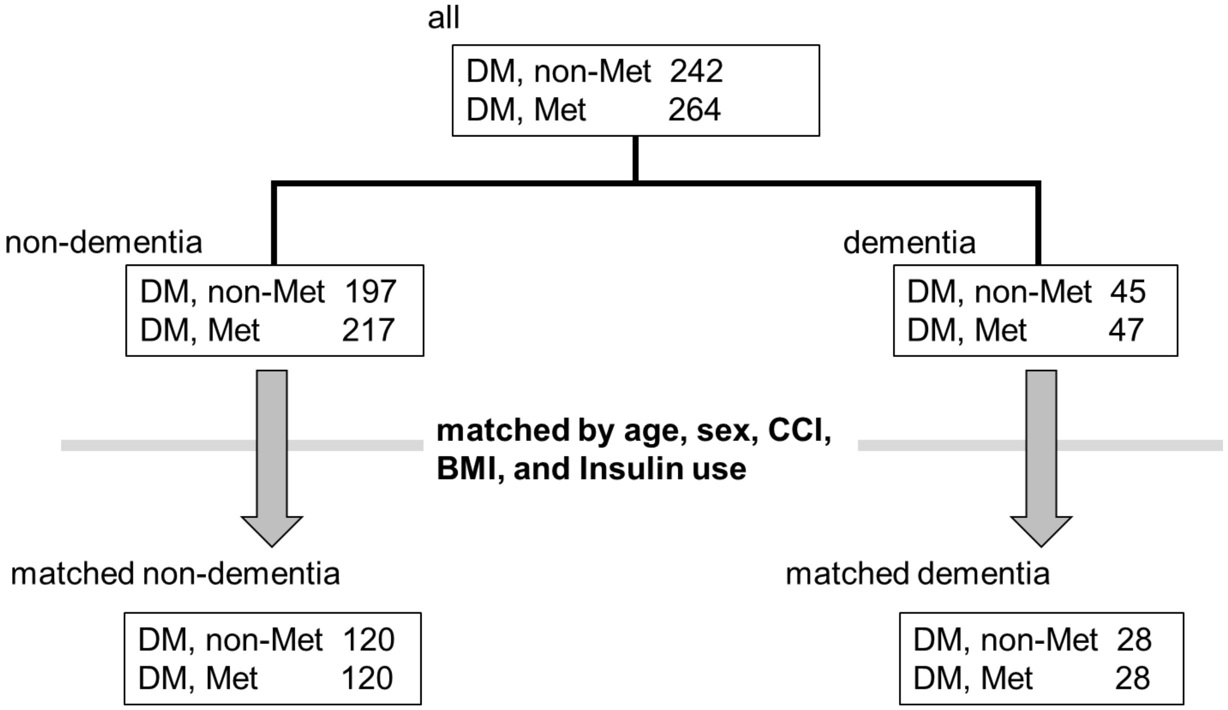


**Supplementary Figure 1**

Propensity score matching process

### Supplementary Figure 2

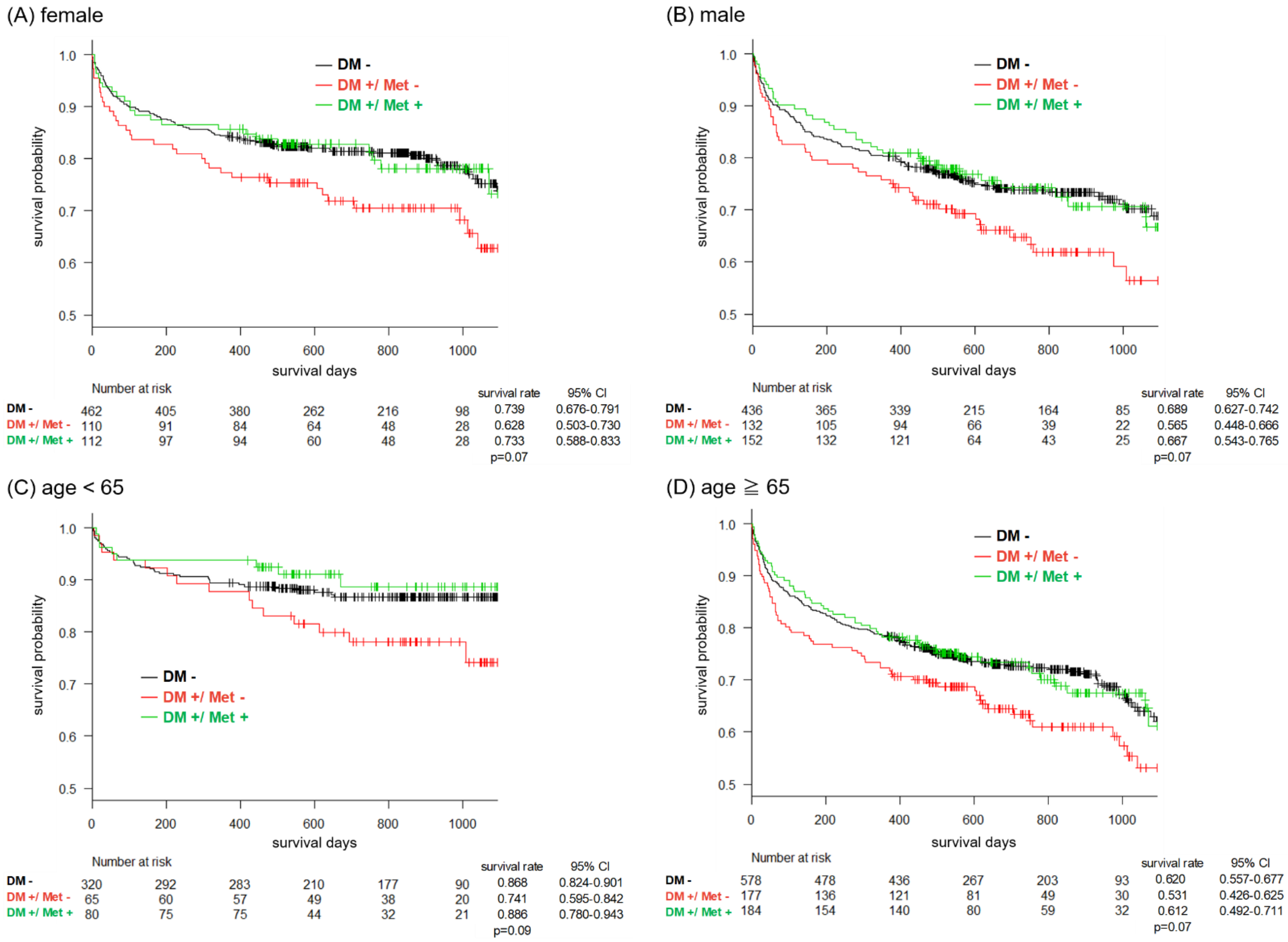


**Supplementary Figure 2**

Kaplan-Meier cumulative survival curve over 3 years based on (A) female only cohort, (B) male only cohort, (C) age <65 years cohort, (D) age ≥65 years cohort.

### Supplementary Figure 3

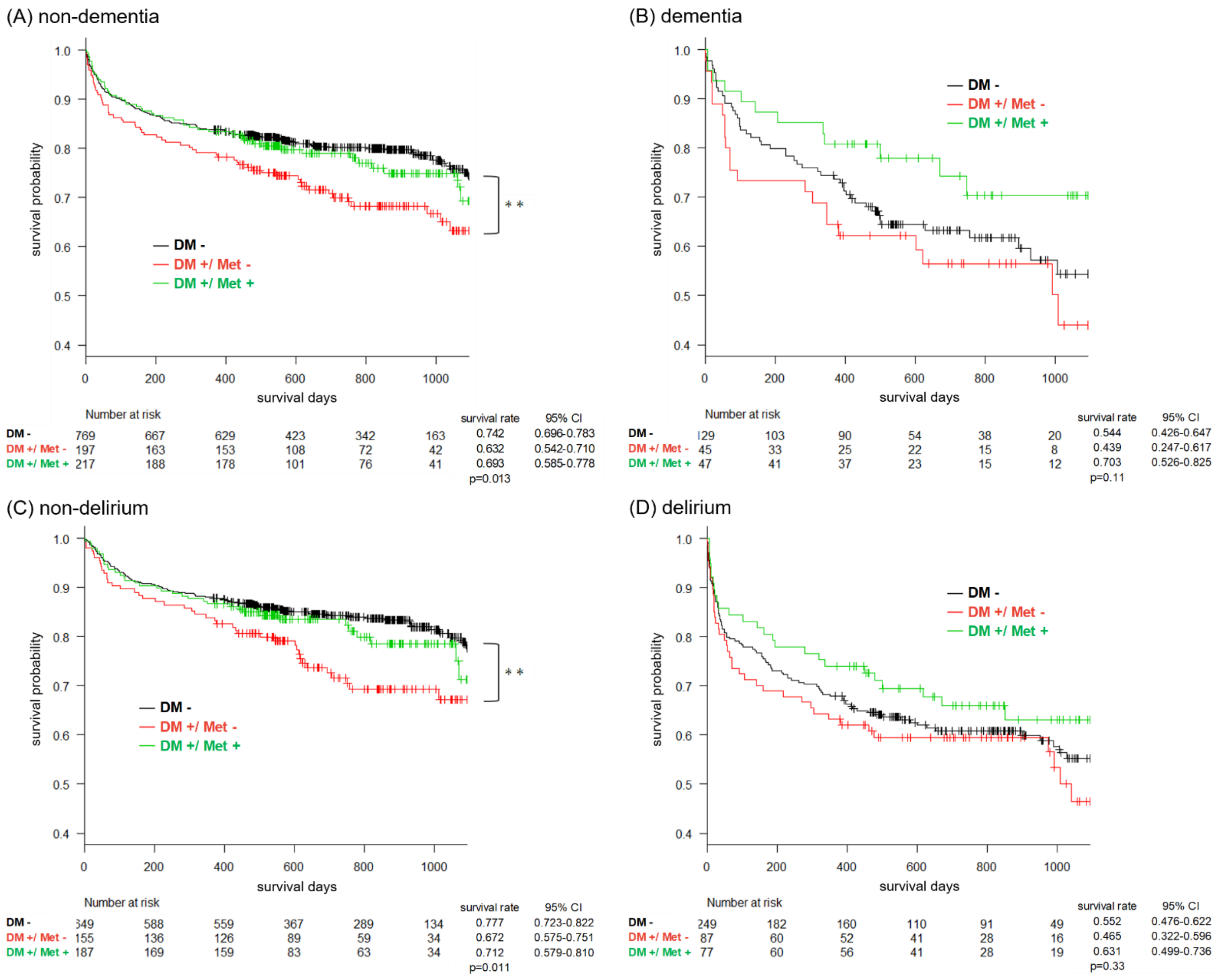


### Supplementary Figure 3

Kaplan-Meier cumulative survival curve over 3 years based on (A) non-dementia cohort, (B) dementia cohort, (C) non-delirium cohort, (D) delirium cohort.

** p<0.01

**Supplementary Figure 4**

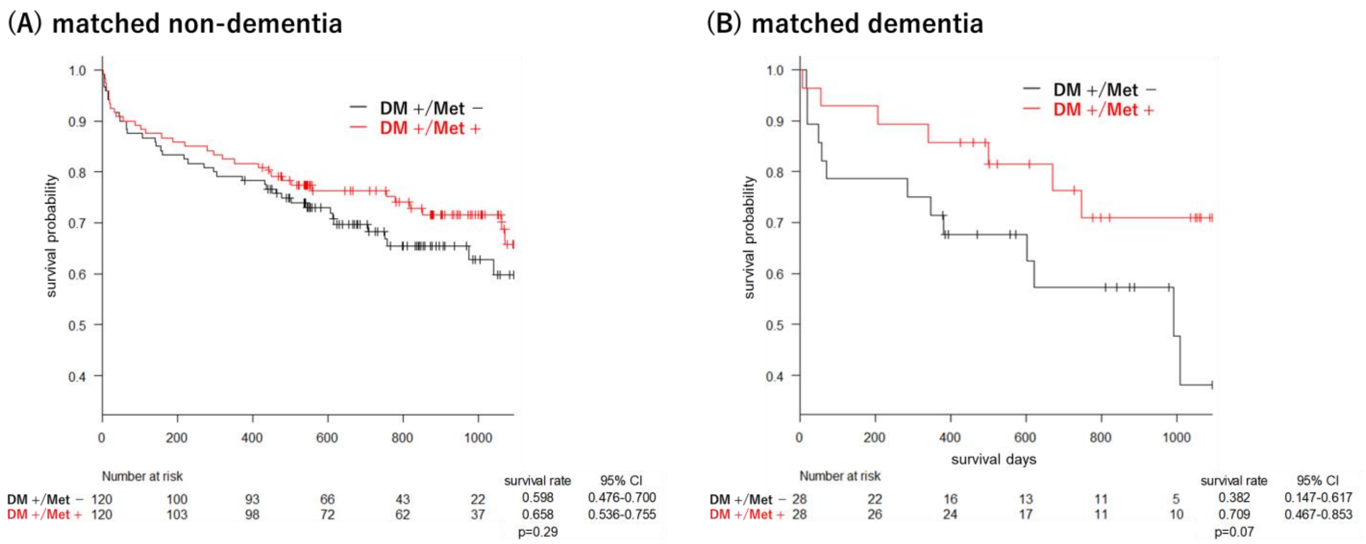


**Supplementary Figure 4**

Kaplan-Meier cumulative survival curve over 3 years based on (A) matched non-dementia cohort and (B) matched dementia cohort.
